# Facilitators and Barriers to the Implementation of the WHO Eight-Contact Antenatal Care Model in Osun State, Nigeria: A Qualitative Study

**DOI:** 10.1101/2025.09.20.25334324

**Authors:** Adebisi Oluwayomi Tomilayo, Akinwaare Margaret

## Abstract

**Background:** The eight-contact antenatal care (ANC) model was introduced by the World Health Organization (WHO) in 2016 to standardize maternal care and reduce maternal and fetal complications by decreasing morbidity and mortality. Despite its widespread adoption, implementation in many low- and middle-income countries, including Nigeria, has been inconsistent. This study explore the factors that facilitate and hinder the implementation of the eight-contact ANC model in specific healthcare facilities in Osun State, Nigeria.

**Methods:** A qualitative descriptive design was adopted, involving in-depth interviews with antenatal care providers and focus group discussions with pregnant women. Purposive sampling was used to select participants. Data were analysed thematically and triangulated with existing literature to enhance credibility and validity.

**Results:** Findings indicated that the eight-contact ANC model has not been widely adopted in the studied facilities, as most health facilities still follow traditional visit schedules based on gestational age. Key barriers include a lack of provider awareness and training, staff shortages, inadequate infrastructure, and financial constraints. Pregnant women also reported challenges such as low health literacy, delayed initiation of care, long travel distances, and negative attitudes from providers. Facilitators of implementing the model include healthcare providers’ willingness to adopt new approaches, availability of educational materials, client motivation regarding fetal well-being, and effective health education during visits.

**Conclusion:** A significant implementation gap exists between WHO guidelines and real ANC practice in Osun State. Addressing systemic weaknesses such as inadequate training, poor supervision, and lack of institutional support is vital. Policy interventions focusing on capacity building, client education, resource provision, and digital health innovations are necessary. Both healthcare providers and pregnant women have shown openness to the eight-contact model, indicating the potential for improved uptake through strategic reforms.

## 1. Introduction

The United Nations (2020) has recognized ANC as a key component of maternal health and a crucial factor in reducing maternal and infant mortality rates globally (Wang et al., 2023). Antenatal care (ANC) is defined as the care received by pregnant women and teenage girls from qualified medical professionals including risk assessment, health promotion, health education, the prevention and treatment of pregnancy-related or coexisting medical conditions; ANC is essential for achieving favorable pregnancy outcomes and ensuring the best possible health for both mother and unborn child during pregnancy (Twagirumukiza, Bubanje et al., 2024). Approximately 287,000 women has been recorded to have died intra and post pregnancy in which SSA has a larger value of 70% of maternal deaths (202,000 out of 287000); these figures indicate a need for prompt action to meet the envisaged SDG goal by 2030 ‘a reduction in maternal mortality to less than 70 per 100,000 live births’ (Lateef, Kuupiel, Mchunu et al., 2024). In low- and middle-income countries, the use of ANC has increased over time; however, this growth has not been associated with a decrease in maternal death or morbidity, thus there is a need for a paradigm change from simply using ANC to using ANC in a quality manner (Dickson, Boateng et al., 2023).

In 2016, the World Health Organization (WHO) increased the recommended number of antenatal care (ANC) visits from four to eight, aiming to reduce maternal mortality and morbidity (Dickson, Boateng et al., 2023). This revision has contributed to the earlier identification and prevention of adverse pregnancy outcomes, reinforcing ANC as a key intervention in healthcare strategies designed to lower maternal and perinatal mortality and morbidity. 2021).Despite the introduction of the 2016 WHO ANC model, which recommends a minimum of eight contacts to replace the previous four-visit approach, maternal and perinatal outcomes in many low- and middle-income countries remain suboptimal (Abdelmola, 2023). In countries like Nigeria, despite the high burden of maternal and neonatal mortality, ANC remains underutilized, often due to poor access, delayed initiation, and insufficient quality of care. Nigeria ranks second globally for newborn deaths and accounts for over 28% of maternal deaths, highlighting the urgent need not only to scale up ANC coverage but also to ensure its delivery meets recommended standards (Abdelmola, 2023; WHO, 2023).

Through different periods the World Health Organization (WHO) has established numerous recommendations and guidelines to enhance healthcare services provided to pregnant women. The antenatal care model established in the US in 1900 emphasized hospital visits instead of quality (Nwabueze, Okeke, Nwevo et al., 2023) but failed to lower maternal mortality rates because of execution challenges and excessive resource usage. Also, the model’s focus on evidence-based therapies, clinical assessments, health education, early detection of complications, and appropriate vaccinations and nutritional supplements has been criticized for not addressing all health needs (WHO, 2023). The 2000s Focused Antenatal Care (FANC) model, linked to more perinatal deaths due to increased stillbirths, has faced criticism for not being comprehensive enough to address all health needs, especially in low-resource settings (Nyumwa, Bula, & Nyondo-Mipando, 2023). FANC model consists of at least four ANC contacts for pregnant women in the case of uncomplicated pregnancies, with the first contact occurring in the first trimester (Kourouma et al., 2021). The eight-contact WHO ANC model emphasizes that a woman’s ‘contact’ with her provider should be more than just a ‘visit,’ but rather an opportunity for high-quality care that includes timely and pertinent information, medical attention, and support throughout the pregnancy (Sserwanja, Musaba et al., 2022). The new WHO ANC guidelines guarantee a more active relationship between pregnant women and their healthcare professionals by referring to “contact” rather than “visit.” This further guarantees that women will interact with the healthcare providers more, providing more chances for health education and birthing preparation (Sserwanja, Musaba et al., 2022).

Antenatal care providers are crucial in delivering ANC services, and their perspectives, experiences, and challenges are key to understanding the facilitators and barriers of the eight-contact model. Antenatal care providers are the most frequently available skilled maternity care providers in rural communities; likewise, pregnant women are the recipients of the care rendered. There is a paucity of research on facilitators and barriers influencing the implementation of the eight-contact antenatal care model focusing in all levels of healthcare, also limited research has been done on key stakeholders, antenatal care providers and pregnant women’s perspectives on eight-contact model using qualitative study in Osun State and Nigeria. Understanding their experiences in maternity care is essential to identify and provide innovations for the implementation of eight contact model, hence the need for this study; this work aims to cover this knowledge gap. The findings are expected to inform the development of evidence-based interventions and recommendations for individuals, educators, and policymakers.

## 2. Methods

### Design and Setting

This study used phenomenological theory for this exploratory qualitative research. Semi-structured interviews and a focus group discussion guide were employed to explore the perspectives of antenatal care providers and pregnant women on the eight-contact ANC model, as well as its facilitators and barriers.

The study was conducted in selected public and private healthcare facilities across Osun State, Nigeria. Osun State is located in the southwestern region of Nigeria, with an estimated population in 2013 of 4,274,858 people (National Population Commission (NPC), 2009). Osun State has three senatorial districts, a total of 30 Local Government Areas, most of which are predominantly rural. There are 736 public health facilities (678 primary, 54 secondary, and 4 tertiary facilities) and 359 private health facilities (Federal Ministry of Health, 2011), providing a representative sample of the state’s health system. Seven hospitals will be selected: Obafemi Awolowo Teaching Hospital Complex, Wesley Guild Hospital, State Hospital Ilesa, Seventh Day Adventist, Ooke-oye Primary Health Care Centre, Aderemi PHC, and Adenle Memorial Hospital. The following facilities will be selected purposively based on the number of pregnant women who access their ANC services, and the above facilities cut across every level of health (Primary, Secondary, Tertiary, and Private facilities).

### Participants and Sampling

Participants included pregnant women and ANC providers selected through purposive sampling to ensure diverse perspectives.

In the first stage, one senatorial district (Osun East) was selected purposively based on the availability of all levels of health care and representative of the state health system. The Osun East consists of two major zones, Ife and Ijesa zones. In the second stage, convenience sampling was used to select two tertiary facilities, two secondary facilities, two primary health centers, and two private health centers from both zones based on the availability of antenatal and delivery services. The health facilities selected are Obafemi Awolowo Teaching Hospital Complex, Wesley Guild Hospital, State Hospital Ilesa, Aderemi Primary Health Care Centre, Oke-ooye PHC, Adenle Memorial Hospital, and Seventh Day Adventist.

From the eight health facilities selected, antenatal care providers who are leaders of each unit were chosen for key-informant interviews based on their official positions and the roles they play in policy formulation and the delivery of maternal health services in the health facilities. Focus group discussions were conducted among pregnant women from the selected health facilities, and pregnant women were chosen using convenience sampling, with each group of 8 pregnant women, until data saturation was reached.

### Data Collection

A key-informant interview guide was used to collect data among leaders of ANC providers, and Focus Group Discussions (FGDs) among pregnant women. The key informant interviews were conducted using the English language and took place in the offices of the providers. The researcher scheduled an interview time and date with the participants at their convenience. The focus of the study and its benefits were explained to the participants. The participants were also informed about voice recording for analysis.

Their permission for recording was taken, and encouraged not to mention their names or the name of their institution during the interview. Notes were taken by the research assistants during the interview, which were used later to compare with transcribed interviews for proper understanding. At the succeeding interview audio recording of the interviews was played to the participants, and the field notes were read to them for confirmation and clarification of their intentions. In addition, it can help in getting more information regarding the discourse. The average duration of the interviews will be 30 minutes. Focus group discussions were conducted in English and Yoruba, same was recorded using a recorder. It was done on clinic days.

### Ethical considerations

Ethical approval was obtained from selected health facilities in Osun State. Ethical clearance was obtained from Obafemi Awolowo Teaching Hospital Complex with protocol number: ERC/2025/04/19 dated 23^rd^ April, 2025, Seventh Day Adventist Hospital with protocol number SERC-2024-11-0041 dated 6^th^ January 2025, and Osun state Ministry of Health dated 7th March 2025.

Informed Consent, confidentiality, and data Security were ensured. Only the research team had access to the password-protected files where the data were safely kept.

### Data Analysis

Thematic analysis was employed to develop a rich thematic description and to analyze patterns of shared meanings of the experiences of the participants, using qualitative data analysis software (NVivo 15 software). Triangulation and member checking were used to ensure trustworthiness.

familiarizing oneself with the data was done by rereading the data and transcribing initial ideas; initial codes were generated by coding interesting features of the data and collating data relevant to new codes. Themes were generated and checked to see if the emerging themes work in relation to the codes. Potential themes were reviewed, defining and naming themes followed, and the report was produced. Data saturation was assessed iteratively during the data collection process. Saturation was deemed to have been reached when successive interview revealed no new codes or themes, and the same patterns began to recur. By the 4^th^ focus group discussion, thematic redundancy was evident, and no novel insights were observed.

## 3. Results

The results were based on one-on-one in-depth interviews with 8 Antenatal Care providers of ages ranging from 34 to 52; six were married, and the other two were widowed. Five participants attained a diploma in education, and three received a degree. All participants are Yoruba by tribe. The ANC providers involve 2 Nurses, 4 Community Health workers, and 2 Medical Doctors. Four focus group discussions were carried out with 8 pregnant women in each focus group aged 16–38 who attended ANC. Shown in Table 1.

**TABLE 1:** Socio-demographic Data of antenatal care providers.

| **ANTENATAL**<br>**CARE**<br>**PROVIDER** | **YEARS**<br>**EXPERIENCE** | **FACILITY**<br>**TYPE** | **EDUCATION**<br>**LEVEL** |
| --- | --- | --- | --- |
| ANC Provider 1 | 10 | Private | Diploma |
| ANC Provider 2 | 9 | Private | MBBS |
| ANC Provider 3 | 9 | Private | Diploma |
| ANC Provider 4 | 7 | Primary | Diploma |
| ANC Provider 5 | 8 | Primary | BSc |
| ANC Provider 6 | 10 | Secondary | BNSc |
| ANC Provider 7 | 4 | Tertiary | MBBS |
| ANC Provider 8 | 5 | Tertiary | Diploma |

### 3.1 Current Status of Implementation

The theme “implementation status” was developed for this section to examine the current status of eight-contact antenatal Care in each selected facility and gain insight into how their antenatal services work. The sub-themes identified under the theme of implementation status include Awareness of the Eight-Contact ANC Model, lack of awareness, and the usability of the eight-contact model.

> *“I am not aware of the implementation of the eight-contact ANC model; perhaps it was just introduced. I have not heard about the model at all*.*” {ANC Provider 1: Private Facility}*
>
> *“I have heard about the eight-contact antenatal model. But our contact with pregnant women depends on the week of their trimester. those in the first trimester to mid-second trimester, we give them 4 weekly appointments, and when they are like 30 weeks, we give them 2 weekly appointments. After that, for those who are like 34 weeks, we give them a weekly appointment*.*” {ANC Provider 3: Private Facility}*
>
> *“We don’t have a particular model that we use in this facility. I could remember there was a training on how often pregnant women should come; it has been few years back, but the number of visits in this facility is dependent on the pregnant woman*.*” {ANC Provider 4: Primary Facility}*
>
> *“I am not aware of the implementation of the eight-contact ANC model; perhaps it was just introduced. I have not heard about the model at all*.*” {ANC Provider 1: Private Facility}*
>
> *“We give appointments based on the gestational age and trimester. We use the traditional model in this facility, and this is basically because the majority of pregnant women tend to miss appointments. By giving them frequent contact, you can make up for the missed appointment. Imagine a booked patient coming in with a stillbirth; to avoid this occurrence, we use traditional antenatal care” {ANC PROVIDER 7: TERTIARY}*

### 3.2 Facilitators of Implementation

Implementing the eight-contact antenatal care model requires facilitators who support pregnant women and their care providers. This theme is represented by four distinct sub-themes: Four distinct categories facilitate implementing the eight-contact antenatal care model among pregnant women and antenatal care providers: Capacity Building and Educational Resources, Policy-Level Support, Health System Strengthening, and Community Awareness and Affordability.

Some healthcare workers expressed openness to adopting new guidelines. Posters, leaflets, and antenatal classes helped promote the model. Women were motivated by concern for fetal well-being and trust in modern healthcare.

> *“Then we can assist in terms of the cost the pregnant women have to spend at each visit by reducing the cost, this will encourage them to come for the scheduled visit”. (ANC PROVIDER 1)*
>
> *“Training of healthcare workers on the latest guidelines in antenatal services will help in implementing any policy that is being introduced” (ANC PROVIDER 4, 5, & 7)*.

Our analysis revealed that, for the eight-contact model to be implemented, policymakers have a significant role to play in enhancing the use of the model by antenatal care providers among pregnant women. The data suggest that policymakers have the potential to transform passive healthcare recipients into active participants in their care journey. This transformation is facilitated by improved access to information, support from policymakers. A healthcare provider’s perspective reinforced this benefit:

> *“This is a private facility, we make use of traditional antenatal care, we have not heard of the eight-contact model, but we can implement it. For this to occur, we need to inform the Medical Director on how to go about the eight-contact ANC model” (ANC PROVIDER 1)*.

### 3.3 Theme 3: Barriers to Implementation

Thematic clusters of these barriers were identified from the patterns and issues. The sub-themes and items as identified by the providers are summarized in the table below.

Table 2 demonstrate that the obstacles to realizing the Eight-Contact ANC model could be divided into four major categories: behavioral, accessibility, health system, and communication. Behavioural barriers like bad attitudes, ignorance and poor compliance, as shown by pregnant women hinder continued ANC participation. Accessibility is impeded by access challenges such as long travel and bad road networks. Challenges within the health system, especially staffing levels and motivation, undermine service delivery, and unmet communication needs, such as insufficient information and branded training, constrain providers’ effectiveness and patients’ participation. Overcoming these barriers is vital for better uptake and implementation of the ANC model.

**Table 2:**
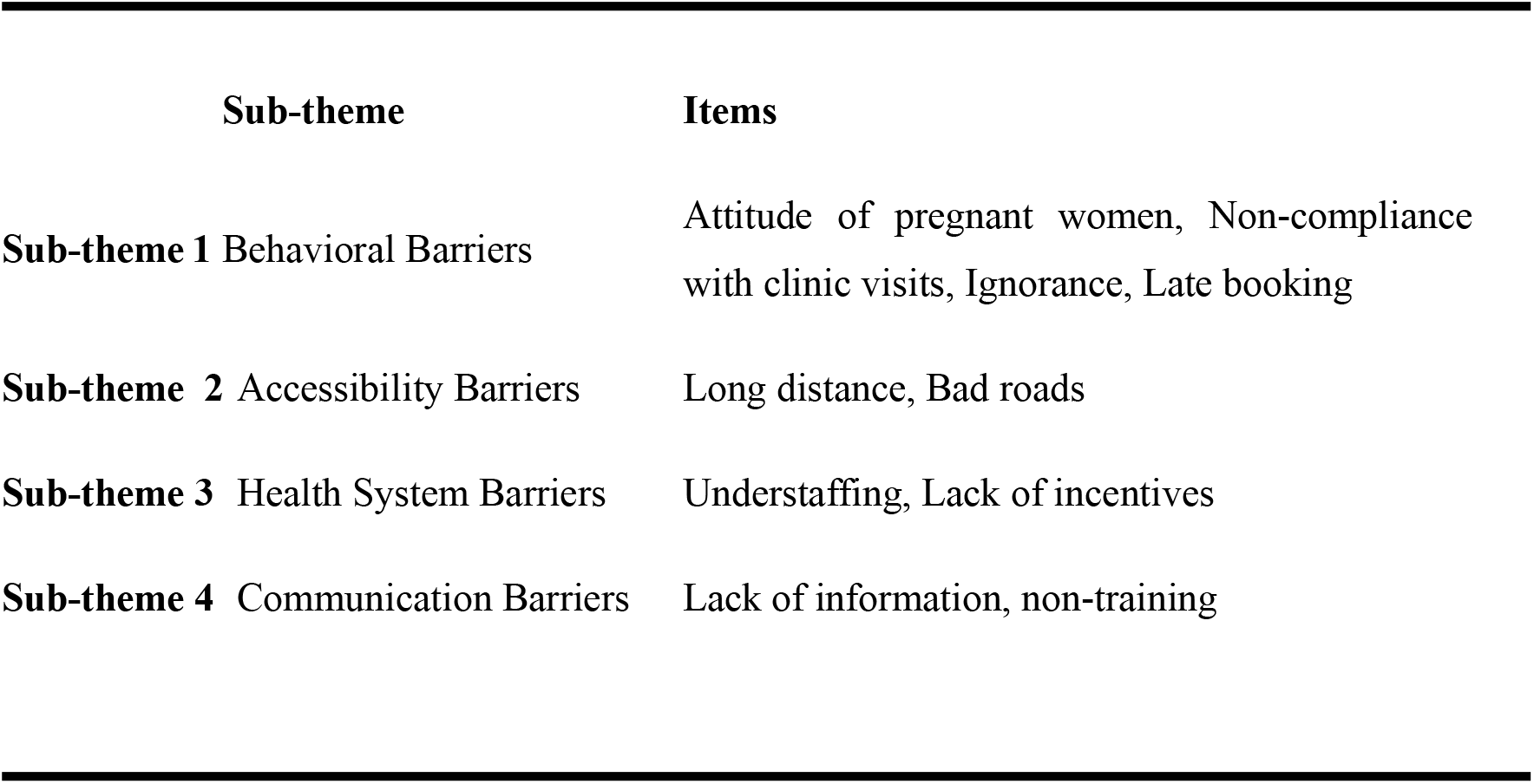
Themes showing barriers to implementation of the Eight-contact antenatal Model.

**TABLE 3:** Facilitators for Implementing the Eight-Contact Antenatal Care Model.

| **Theme 1: Facilitators for Implementing the Eight-Contact Antenatal Care Model** |  |  |
| --- | --- | --- |
| **Cluster** | **Items** | **Sub-theme** |
| **Cluster 1** | - Training - Availability of fliers and materials | **Capacity Building and Educational Resources** |
| **Cluster 2** | - Informing the policymaker | **Policy-Level Support** |
| **Cluster 3** | - Enough staffing – Funding | **Health System Strengthening** |
| **Cluster 4** | - Cost reduction - Educating pregnant women | **Community Awareness and Affordability** |

> *“ANC Provider 7: Attitude of pregnant women……. ANC Provider 1: Attitude of pregnant women,……. ANC Provider 8: Staffing I’ll say pregnant women’s attitude*.*”*.

Provider 5 mentioned that some women travel long distances, which discourages regular visits.

> *“Some pregnant women come from a far distance, so they do not adhere*…*”*

Bad Roads were cited by Provider 1; poor road conditions further hinder access.

> *“The road leading to our facility is bad*…*”*

### 3.4 Theme 4: Opinion of Pregnant Women on the Implementation of the Eight-Contact Model

A General Theme was developed: the opinion of pregnant women. 3 Subthemes were identified, which are Low Awareness of the Eight-Contact ANC Recommendation, perceived Benefits of Frequent ANC Visits, Barriers/motivators to ANC Attendance. Fig 1.0 displayed the subthemes and pregnant women’s opinions.

Across all four FGDs, it was consistently found that none of the 32 women were aware of the WHO recommendation to attend at least eight ANC visits during pregnancy. Almost every participant echoed the statement, *“I have not heard about that*.*”* This indicates a critical information gap regarding global health guidelines and the need for enhanced health education and communication from healthcare providers.

Despite the lack of awareness, most participants expressed positive attitudes toward the concept of frequent ANC visits once it was introduced to them. The perceived benefits include:

- Monitoring baby’s health: *“It will enable us to check on our baby’s welfare*.*”*
- Monitoring maternal health: *“It will give room for monitoring your blood pressure*.*”*
- Adequate care and early detection of complications: *“Ensures care for myself and the baby,” “During pregnancy, anything can go wrong*.*”*

Some participants travel long distances to reach the clinic, making regular visits burdensome.

> *“I live in a far place from this place, so I can’t come frequently*.*”*

Instances of rude behavior from healthcare staff discouraged attendance.

> *“The way she attended to me was rude, and I have decided I won’t go back for the test*.*”*

Participants also identified systemic issues, such as staff Shortages, several women noted that student nurses were often responsible for care, possibly affecting service quality. Long Waiting Times were consistently raised as a major issue. Inadequate Infrastructure and Resources indirectly reduce women’s willingness to return or complete the eight recommended visits.

## 4. Discussion

Findings highlight a clear discrepancy between WHO recommendations and actual ANC delivery.

### Theme 1: Implementation Status

The WHO’s 2016 eight-contact ANC model was designed to improve maternal and fetal outcomes through increased frequency and quality of contact. However, findings from this study in Osun State, Nigeria, revealed poor adoption of the model, with most facilities still using traditional ANC schedules based on gestational age rather than structured contact points. While some providers reported that women attended more than eight visits, these were unstructured and undocumented, aligning with findings from Mengistie (2024) in Ethiopia and Ekholuenetale et al. (2021) across LMICs. A widespread lack of awareness and training about the eight-contact model was evident among providers, pointing to gaps in policy dissemination and institutional support. Despite national statistics showing low ANC contact rates (Ekholuenetale et al., 2020), some areas in Osun State reported higher figures (Fagbamigbe et al., 2018), though actual model compliance remained questionable.

### Theme 2: Facilitators of Implementation

Key facilitators included provider training, availability of educational materials (calendars, handbooks), and client motivation. Training enhances provider competency and communication (Khatri et al., 2021), while structured materials improve care quality (Owili et al., 2019). Cost reduction was identified as critical for improving ANC attendance, consistent with findings by Atuhaire et al. (2019) linking higher income to better ANC compliance. Institutional support, especially from policymakers and health administrators, was emphasized as vital to operationalizing the model, echoing Kourouma et al. (2021) on the importance of governance. Adequate staffing and facility readiness further enhanced service quality, while community education and the use of digital health tools were seen as effective strategies to improve model uptake (Alibhai et al., 2022; Wulandari & Laksono, 2020).

### Theme 3: Barriers to Implementation

Financial hardship was the most prominent barrier, with economic status strongly affecting ANC compliance (Ekholuenetale et al., 2021). Late booking, missed appointments, and limited awareness among pregnant women reflected broader issues of health literacy (Mulondo, 2020; Oguntunde et al., 2019). Distance to health facilities and poor road infrastructure disproportionately affected rural dwellers (Tessema et al., 2020; Dickson et al., 2023). Staffing shortages, provider burnout, and lack of professional motivation were major institutional challenges (Khatri et al., 2022; Seyoum et al., 2021), compounded by outdated training and a lack of supervision or policy enforcement (Tessema et al., 2021). The absence of standard protocols and monitoring tools hindered consistent delivery of the eight-contact model across facilities.

### Theme 4: Pregnant Women’s Perspectives

Focus group discussions revealed a widespread lack of awareness of the eight-contact model among pregnant women. However, upon learning about its purpose, women expressed positive attitudes and acknowledged the benefits of regular ANC attendance for fetal well-being, aligning with findings by Ali et al. (2025) and Rahman et al. (2025). Barriers such as cost, long wait times, and negative staff behavior were significant deterrents to attendance, echoing findings from Jitu et al. (2025), Andiso et al. (2024), and Mlambo & Ibeziako (2025). Despite these challenges, motivators such as concern for the baby, quality of information received, and respectful care encouraged repeat visits. Notably, satisfaction with previous ANC services sometimes led to discontinuation, suggesting that context-specific factors influence adherence (Abera et al., 2023).

### 5.1 Summary of Findings

This study examined the implementation of the WHO-recommended eight-contact antenatal care (ANC) model in selected public and private healthcare facilities in Osun State, Nigeria. Despite WHO’s 2016 guidelines aimed at enhancing maternal and fetal health through structured and frequent contacts, the findings revealed limited adherence. Most facilities still follow traditional models, scheduling visits based on symptoms rather than proactive planning. Many providers were unaware of or untrained in the eight-contact model, resulting in inconsistent application. Likewise, pregnant women showed limited awareness, though they responded positively when educated about its benefits.

Barriers to full implementation included lack of training, poor infrastructure, financial constraints, negative staff attitudes, long wait times, and poor transport access. However, key facilitators identified were client motivation driven by fetal health concerns, positive ANC experiences, and the use of educational materials and structured health talks.

### 5.2 Conclusion

This study concludes that Osun State has not effectively implemented the eight-contact ANC model due to systemic challenges such as insufficient provider training, weak institutional support, and socioeconomic limitations among clients. Although women value ANC services, gaps in awareness, structural constraints, and negative care experiences hinder full utilization. A disconnect remains between WHO guidelines and real-world clinical practice in these settings.

#### Limitations of the Study

1. The research focused on selected health care facilities in Osun State, which may not represent the whole state or other states in Nigeria; however, the study carefully selected the health facilities to ensure good representation.
2. While the qualitative approach offered rich insights, there were no quantitative outcome measures incorporated, such as a proportionate increase in ANC compliance over time, which might restrict capturing the true effectiveness of the implementations

### 5.3 Recommendations

To bridge this gap, the following actionable recommendations are proposed:

1. Provider Training, regular in-service training and refresher courses on WHO ANC guidelines should be institutionalized.
2. Health facilities should develop and distribute standard operating procedures, calendars, and checklists to promote consistency.
3. Local outreach through media and community health leaders is essential to raise awareness and promote attendance.
4. Policymakers should consider subsidies or conditional cash transfers for pregnant women from low-income households.
5. Stronger supervision mechanisms are needed to enforce compliance with the eight-contact model.

### 5.4 Contribution to Knowledge

This study adds context-specific insights into the barriers and facilitators influencing the eight-contact ANC model in a Nigerian setting. Key contributions include:

- Highlighting the socio-cultural and economic dimensions that affect ANC adherence.
- Integrating perspectives from both healthcare providers and pregnant women.
- Revealing a disconnect between global policy and local clinical practice.
- Providing a foundation for designing targeted, culturally relevant interventions.

### 5.5 Implications for Nursing

#### Policy

The study emphasizes the need for national nursing policies that incorporate the eight-contact ANC model into framework, alongside resource allocation for training and SOP development.

#### Practice

Nurses and midwives must shift from traditional care to a structured, quality-driven ANC model, ensuring documentation and respectful, culturally sensitive care.

#### Research

Longitudinal and interdisciplinary nursing studies are needed to evaluate how changes in training, policy, or financial incentives impact ANC adherence and outcomes.

#### Education

Nursing curricula should be revised to reflect current WHO ANC guidelines, with an emphasis on clinical simulations, systems thinking, and public health leadership.

## Data Availability

All relevant data are within the manuscript and its Supporting Information files.

## 7. Acknowledgments

The author thanks all antenatal care providers, pregnant women who made themselves available for the research, and my research supervisor, who contributed immensely to this study

## Data Availability

All relevant data are within the manuscript. Transcripts and anonymized interview data that support the findings of this study are available from the corresponding author upon reasonable request, with appropriate ethical approval.

**Fig 2.**
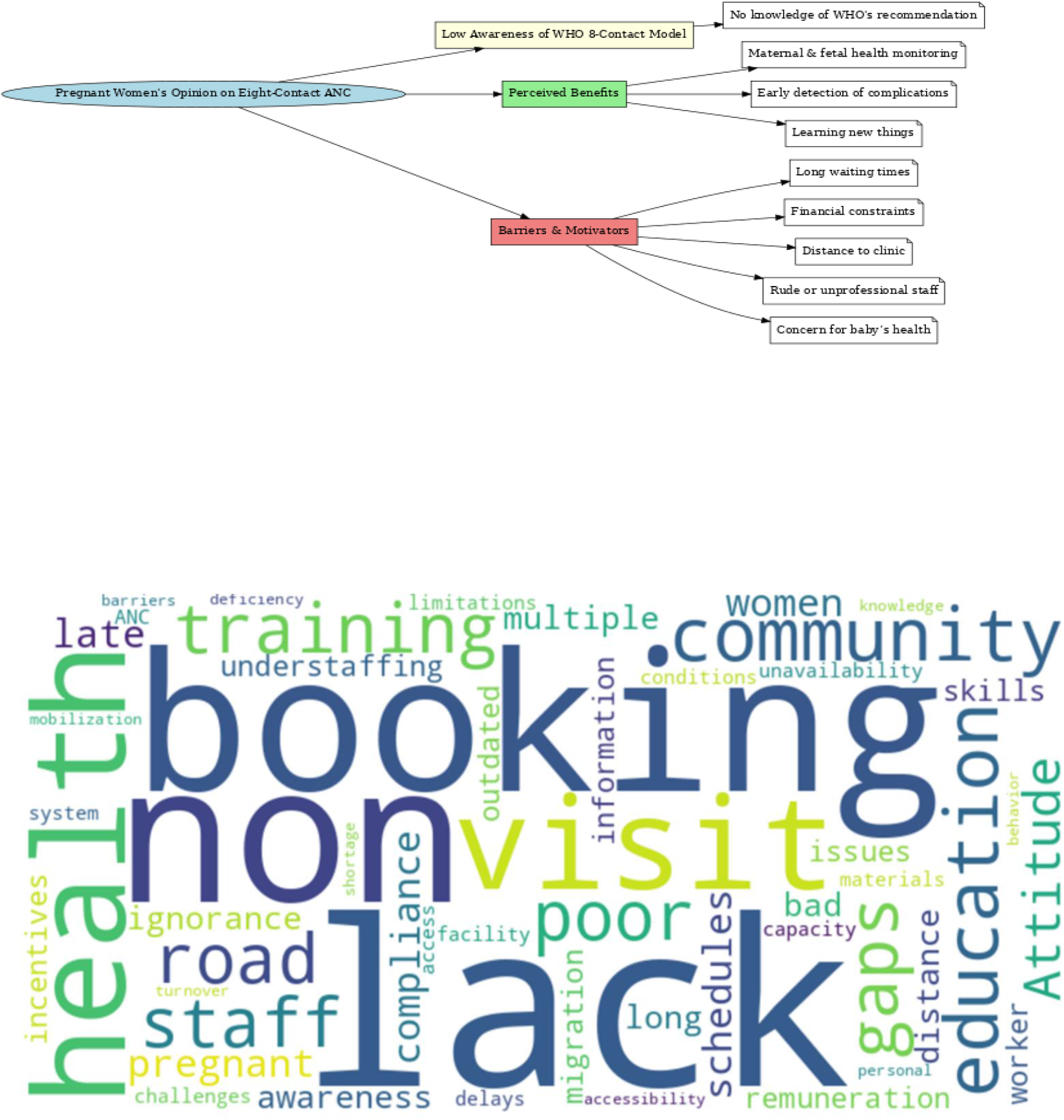
Barriers to the implementation of the eight-contact antenatal model.

